# Data-Driven Development of a Small-Area COVID-19 Vulnerability Index for the United States

**DOI:** 10.1101/2020.08.17.20176248

**Authors:** Ofer Amram, Solmaz Amiri, Emily L. Thorn, Jacob J. Mansfield, Pranav Mellacheruvu, Pablo Monsivais

## Abstract

As the COVID-19 pandemic continues to surge in the United States, it has become clear that infection risk is higher in certain populations, particularly socially and economically marginalized groups. Social risk factors, together with other demographic and community characteristics, may reveal local variations and inequities in COVID risk that could be useful for targeting testing and interventions. Yet to date, rates of infection and estimations of COVID risk are typically reported at the county and state level. In this study we develop a small area vulnerability index based on publicly-available sociodemographic data and 668,428 COVID diagnoses reported in 4,803 ZIP codes in the United States (15% of all ZIP codes). The outcome was COVID-19 diagnosis rates per 100,000 people by ZIP code. Explanatory variables included sociodemographic characteristics obtained from the 2018 American Community Survey 5-year estimates. Bayesian multivariable techniques were used to capture complexities of spatial data and spatial autocorrelation and identify individual risk factors and derive their respective weights in the index. COVID-19 diagnosis rates varied from zero to 29,508 per 100,000 people. The final vulnerability index showed that higher population density, higher percentage of noninsured, nonwhite race and Hispanic ethnicity were positively associated with COVID-19 diagnosis rates. Our findings indicate disproportionate risk of COVID-19 infection among some populations and validate and expand understanding of these inequities, integrating several risk factors into a summary index reflecting composite vulnerability to infection. This index can provide local public health and other agencies with evidence-based metrics of COVID risk at a geographical scale that has not been previously available to most US communities.

## INTRODUCTION

In June 2020, COVID-19 cases in the United States surpassed two million, with an estimated 120,000 associated deaths.^1^ As the global pandemic continues to rapidly expand throughout the country, public health, state, and federal agencies have mobilized to mitigate further spread, however coordination of these efforts remains a challenge.

Research and surveillance data indicate that COVID-19 infection risk is not evenly distributed, but rather increased in certain sociodemographic populations, particularly those who are socially and economically marginalized. Black, Indigenous, and Latinx individuals are at sharply elevated risk of infection compared to white counterparts across numerous U.S. states.^2–4^ In a study of Illinois, Black residents accounted for a striking 37% of confirmed cases, despite comprising just 16% of the state’s population.^5^ Similar disparities have been reported in more granular analysis of urban settings. In New York City, Black and Latinx residents accounted for a greater relative proportion of confirmed cases, yet were less likely to have access to rapid testing.^2,6,7^ Notably, Black, Indigenous, and Latinx individuals are more likely to be essential workers, live in confined housing conditions, and lack adequate healthcare access, however the degree to which these and other factors potentiated by systemic racism may contribute to higher infection risk is not fully known.^5,8^

Assessments of social and economic disparities in infection risk have been limited to state, county, and isolated ZIP code-level analyses. To our knowledge, there has been no national assessment of independent predictors of COVID-19 case burden in the U.S. at the ZIP code level. This information remains critical to the appropriate prioritization of response efforts and allocation of resources both regionally and nationally.

In this study, we assessed socio-demographic and economic predictors of COVID-19 infection risk at the U.S. ZIP-code level, based on sampling of publicly-available data. Findings may support the development of a national vulnerability index and inform a more equitable and effectively coordinated public health response.

## METHODS

### Data

We manually searched for ZIP code level COVID-19 data reported by health authorities. These health authorities have set up dashboards and continuously update and provide reliable and timely data on COVID-19 cases within their jurisdiction. Dashboards listed the number and/or rates of COVID-19 cases by ZIP code. Socio-demographic characteristics of ZIP codes were obtained from the 2018 American Community Survey 5-year estimates. Use of de-identified publicly available data did not warrant a review by an Institutional Review Board.

### Measures

The outcome was COVID-19 diagnosis rates per 100,000 people by ZIP code. We used logarithmic transformation of diagnosis rates for analysis. Explanatory variable included: a) percent of nonwhite population (i.e. non-Hispanic African American, non-Hispanic American Indian or Alaska Native, non-Hispanic Asian or other Pacific Islander, and other non-Hispanic); b) percent of Hispanic population; c) percent of uninsured population; d) percent of population aged 20 to 39; and e) population density. We used logarithmic transformation of population density for analysis.

Associations between the following variables and COVID-19 diagnosis were explored but not included in the multivariable model: a) percent of essential workers including transportation, protective service (firefighting and prevention, and other protective service workers including supervisors and Law enforcement workers including supervisors), health care practitioners, healthcare technical occupations and support; b) percent of African Americans,; and c) percent of population with income below federal poverty line.

### Statistical Analysis

Univariate analysis described measures of central tendency and variability for the explanatory and outcome variables. We used bivariate generalized linear models (GLM) to explore the associations between socio-demographic characteristics of ZIP codes and COVID-19 infection rates. As our goal was to create a ZIP code level vulnerability index across the US from only selected states/jurisdictions where data is available. In order to make sure that the predictor variables selected for the final model had a minimum level of consistency in association with COVID-19 infection across states in our sample. To be considered in the final index, variables had to show association with COVID-19 infection in a minimum of 50% of ZIP codes significantly associated in the same direction and with no more than 15% significant in the opposite direction.

Multivariable models used Bayesian techniques,^9–11^ which are flexible in capturing complexities of spatial data and spatial autocorrelation, the tendency for areas closer to each other to have more similar characteristics than areas farther apart. Mean for factors associated with COVID-19 infection were estimated using Gaussian models. The spatially structured residual was modelled using an intrinsic conditional autoregressive model and the unstructured residual using exchangeability among ZIP codes. The neighborhood adjacency matrix was defined using a U.S. ZIP code shapefile. Models included fixed effects for percent of nonwhite population, percent of Hispanic population, percent of non-insured population, percent of population aged 20 to 39, and population density.

Bayesian models present posterior estimates of the effect of explanatory variables expressed as mean, 95% credible intervals (CI) for the mean, and relative rates (based on the exponentiated crude coefficient). Credible intervals that exclude zero for the mean (or one for relative rates) were considered significant The Integrated Nested Laplace Approximation (INLA) package in R was used for generating Bayesian statistics.

## RESULTS

Of the 32,201 ZIP codes in the U.S., 4,803 ZIP codes from 19 states (15% of all ZIP codes) were included in the analyses (N cases = 668,428). COVID-19 diagnosis rates varied from zero to 29,508 per 100,000 people. Median percentage of non-white population was 31%, percentage of Hispanics 7%, percentage of non-insured population 9%, and percentage of population aged 20 to 39 was 12%. Population density ranged from 0.11 to 111,330 per square miles. Of the total number of ZIP codes included, 3495 (73%) were urban, 1,308 (27%) were small, large or isolated rural areas.

### Variable Selection

Table 1 shows the results of the bivariate GLM model between cases per population and each of the potential predictor for each state. Each of the candidate variables was associated with COVID-19 infection in at least six of the 19 states. However, the direction of association was not always consistent between states for a given variable. For example, essential workers were significant in the same direction in three states and in the other direction in three other states. Based on our inclusion criteria described above, the following variables were included in the final index: Population density, percent uninsured, percent nonwhite, percent Hispanic and percent population aged 20 to 39 years. Although, black race did meet our criteria, we decided to not to include in our final index but instead included non-white race as it was more consistent across all twenty states and it also include other racial categories.

**Table 1.** Univariate generalized linear regression analysis of sociodemographic predictors of COVID-19 infection by state.

| State (n=ZIP codes) | Population Density<br>$\beta$ (SE) | Essential Worker<br>$\beta$ (SE) | Income Below FPL<br>$\beta$ (SE) | Non-white Race<br>$\beta$ (SE) | Black Race<br>$\beta$ (SE) | Hispanic Ethnicity<br>$\beta$ (SE) | No Health Insurance<br>$\beta$ (SE) | Age 20-39 years<br>$\beta$ (SE) |
| --- | --- | --- | --- | --- | --- | --- | --- | --- |
| Arizona (227) | **0.53 (0.04)\*** | 0.01 (0.02) | 0.03 (0.02) | **0.03 (0.01)\*** | **0.31 (0.04)\*** | **0.03 (0.01)\*** | 0.01 (0.02) | **0.11 (0.01)\*** |
| California (155) | **0.08 (0.03)\*** | 0.01 (0.02) | **0.02 (0.01)\*** | **0.02 (0.00)\*** | 0.02 (0.01) | **0.02 (0.00)\*** | **0.04 (0.01)\*** | 0.01 (0.00) |
| Florida (801) | **0.26 (0.02)\*** | **-0.03 (0.01)\*** | **0.02 (0.01)\*** | **0.03 (0.00)\*** | **0.02 (0.00)\*** | **0.02 (0.00)\*** | **0.05 (0.01)\*** | **0.03 (0.01)\*** |
| Georgia (30) | -0.01 (0.35) | 0.02 (0.03) | **0.06 (0.02)\*** | **0.01 (0.01)\*** | 0.00 (0.01) | 0.02 (0.01) | **0.07 (0.02)\*** | -0.02 (0.02) |
| Illinois (498) | **0.25 (0.02)\*** | 0.02 (0.01) | **0.02 (0.01)\*** | **0.02 (0.00)\*** | **0.01 (0.00)\*** | **0.03 (0.00)\*** | **0.11 (0.01)\*** | **0.01 (0.01)\*** |
| Indiana (12) | 0.28 (0.27) | -0.14 (0.07) | 0.02 (0.02) | **0.02 (0.01)\*** | 0.03 (0.01) | 0.04 (0.02) | 0.02 (0.05) | 0.02 (0.04) |
| Iowa (28) | **0.69 (0.30)\*** | -0.24 (0.16) | 0.04 (0.07) | **0.17 (0.06)\*** | **0.22 (0.09)\*** | **0.72 (0.28)\*** | 0.46 (0.31) | **0.18 (0.08)\*** |
| Kansas (11) | 1.20 (0.57) | -0.18 (0.12) | 0.08 (0.06) | **0.06 (0.02)\*** | 0.07 (0.04) | 0.05 (0.03) | 0.09 (0.05) | 0.03 (0.13) |
| Maryland (281) | **0.18 (0.03)\*** | 0.02 (0.01) | **0.02 (0.01)\*** | **0.02 (0.00)\*** | **0.02 (0.00)\*** | **0.05 (0.00)\*** | **0.11 (0.00)\*** | **0.04 (0.01)\*** |
| Michigan (19) | 0.25 (0.026) | **0.28 (0.13)\*** | **0.08 (0.03)\*** | **0.05 (0.02)\*** | **0.08 (0.03)\*** | **0.50 (0.19)\*** | 0.38 (0.23) | **0.06 (0.06)\*** |
| New York (175) | **-0.22 (0.04)\*** | **0.05 (0.00)\*** | **0.01 (0.00)\*** | **0.01 (0.00)\*** | **0.01 (0.00)\*** | **0.01 (0.00)\*** | **0.04 (0.01)\*** | **-0.03 (0.00)\*** |
| N. Carolina (683) | 0.02 (0.03) | **0.04 (0.01)\*** | **0.03 (0.01)\*** | **0.02 (0.00)\*** | **0.02 (0.00)\*** | **0.06 (0.01)\*** | **0.05 (0.01)\*** | 0.01 (0.01) |
| Oklahoma (347) | -0.01 (0.03) | **-0.04 (0.01)\*** | 0.00 (0.01) | **0.01 (0.00)\*** | 0.00 (0.01) | **0.02 (0.01)\*** | 0.01 (0.01) | 0.00 (0.01) |
| Oregon (162) | **0.69 (0.06)\*** | -0.09 (0.05) | -0.01 (0.03) | **0.12 (0.01)\*** | **0.37 (0.08)\*** | **0.13 (0.02)\*** | 0.05 (0.06) | **0.21 (0.02)\*** |
| Rhode Island (75) | **0.60 (0.18)\*** | -0.04 (0.03) | 0.07 (0.04) | **0.06 (0.01)\*** | **0.28 (0.06)\*** | **0.07 (0.02)\*** | **0.27 (0.09)\*** | 0.06 (0.04) |
| S. Carolina (382) | 0.00 (0.03) | 0.01 (0.01) | **0.01 (0.00)\*** | **0.01 (0.00)\*** | **0.01 (0.00)\*** | 0.01 (0.01) | **0.02 (0.01)\*** | 0.01 (0.01) |
| Texas (150) | **0.72 (0.13)\*** | 0.03 (0.03) | **0.07 (0.01)\*** | **0.03 (0.00)\*** | **0.03 (0.01)\*** | **0.02 (0.01)\*** | **0.06 (0.01)\*** | **0.07 (0.02)\*** |
| Virginia (636) | **0.59 (0.05)\*** | 0.00 (0.01) | **-0.03 (0.01)\*** | **0.06 (0.01)\*** | **0.04 (0.01)\*** | **0.17 (0.01)\*** | **0.05 (0.02)\*** | **0.05 (0.01)\*** |
| Washington (131) | **0.52 (0.05)\*** | **-0.13 (0.03)\*** | **0.03 (0.02)\*** | **0.05 (0.01)\*** | **0.12 (0.02)\*** | **0.14 (0.02)\*** | 0.06 (0.05) | **0.06 (0.01)\*** |

\*Univariate regression analysis significant at the $\alpha=0.05$ level, $\beta$ = Beta-coefficient, SE = Standard Error, FPL = Federal Poverty Level

According to INLA models, higher population density, higher percentage of noninsured, nonwhite race and Hispanic ethnicity were positively associated with COVID-19 diagnosis rates (Table 2). We used the coefficients from the INLA mode to create an index that reflected risk of COVID-19 diagnosis rates for ZIP codes across the U.S. The median value for index was 41.9 with an interquartile of 30.3 to 55.4. The distribution of index values are shown in Figure 1 and an illustration of the variation in index values across ZIP codes in Washington are shown in Figure 2.

**Table 2.** Posterior estimates of factors associated with COVID-19 diagnosis rates from the Integrated Nested Laplace Approximation (INLA) models.

| Variables | Mean | SD | 2.5% CI | 97.5% CI |
| --- | --- | --- | --- | --- |
| Population density (log transformed) | 0.19 | 0.01 | 0.16 | 0.21 |
| Percent noninsured | 0.01 | 0.01 | 0.01 | 0.02 |
| Percent nonwhite | 0.03 | 0.001 | 0.03 | 0.03 |
| Percent Hispanic | 0.01 | 0.002 | 0.01 | 0.01 |
| Percent population aged 20 to 39 | 0.001 | 0.005 | -0.01 | 0.01 |
| *Random effects* |  |  |  |  |
| Spatial structure (iCAR) | 0.82 | 0.16 | 0.54 | 1.18 |
| Spatial unstructured (exchangeable) | 3.59 | 0.59 | 2.60 | 4.90 |

**Figure 1.**
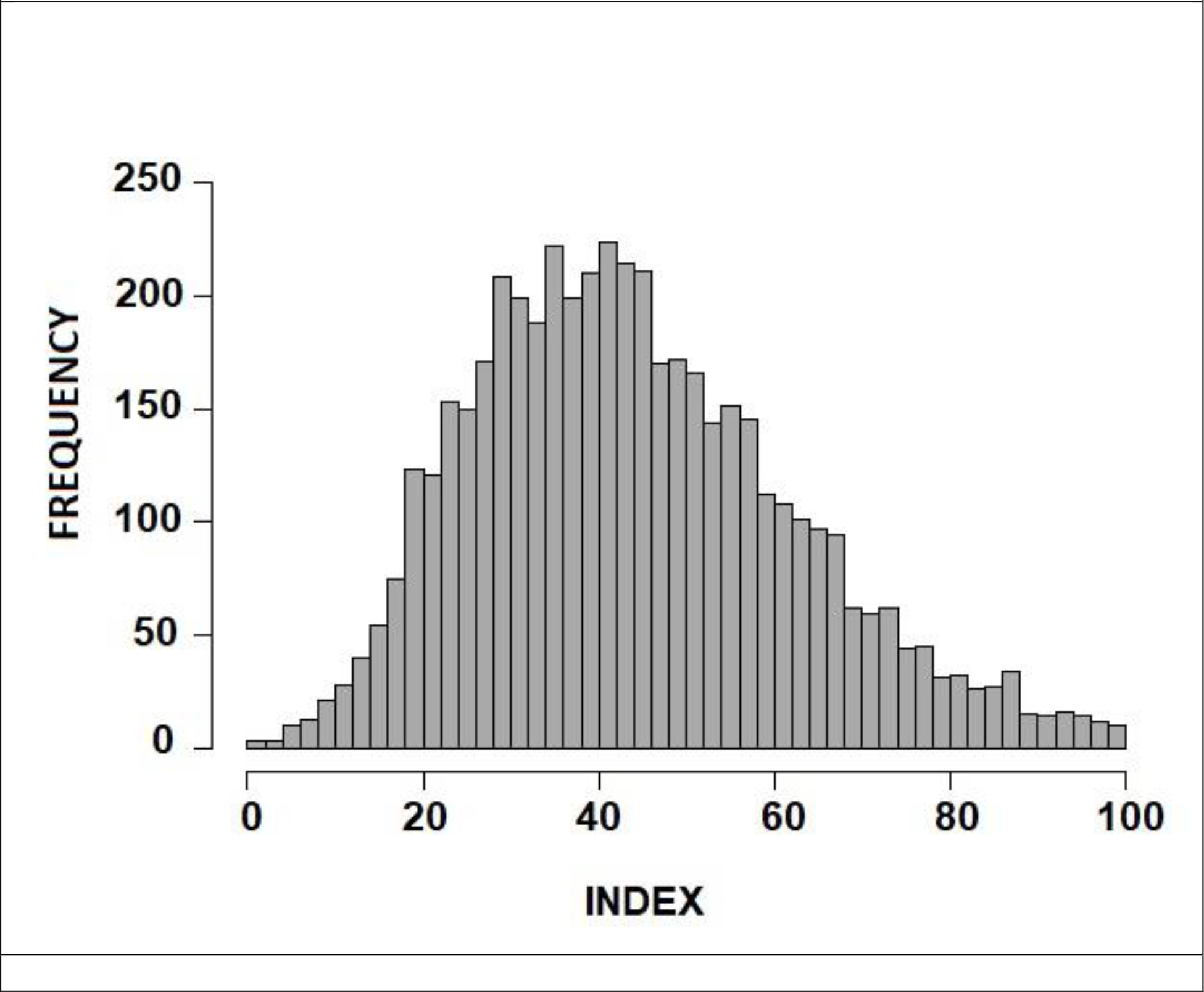
Histogram illustrating the distribution of COVID-19 vulnerability index values for 32,201 ZIP codes in the United States

**Figure 2.**
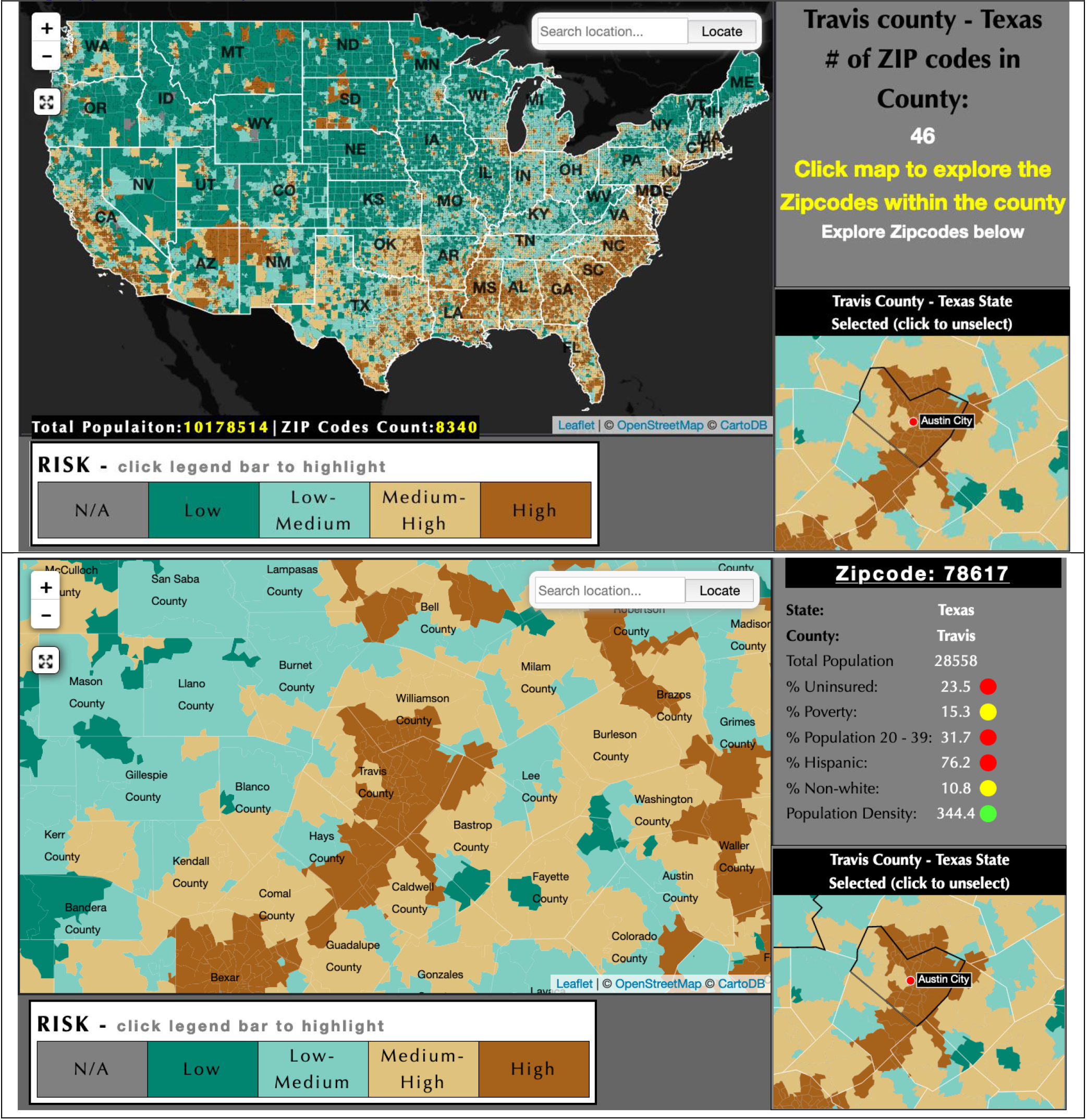
Screen grab of interactive data visualization of COVID-19 vulnerability across the United States (upper panel) and with Travis County, Texas selected for higher-resolution display of ZIP code-level vulnerability in that county (lower panel). Interactive data visualization of index is available here: https://chaselab.net/USACovidIndex/Index.htm

## DISCUSSION

As COVID-19 cases surge in a majority of states, development of a targeted, coordinated public health response is urgently needed. County- and state-level analyses have described local factors modifying risk of infection, including race, ethnicity, occupation, population density, insurance status, and age, however independent predictors of infection remain unknown. In this study, we develop the first data-driven national COVID-19 risk index at the zip-code level, identifying several significant independent risk factors to infection, including race, ethnicity, insurance status, and population density.

### Relation to Previous Studies

Previous studies have consistently identified disparities in COVID-19 case burden by race and ethnicity.^2,4,12^ This study reinforces these findings, where non-white race was associated with a consistently higher risk of infection across all states sampled and both non-white race and Hispanic ethnicity were found to increase risk when controlling for other predictors. Furthermore, though non-Hispanic Black race did not meet the criteria for model inclusion, this population demonstrated increased odds of infection in univariate modeling, with a significant association found in 18 of the 19 states sampled. This further establishes that although multiple communities of color appear to have increased risk of infection, it remains critical to identify and target the unique structural barriers facing Black Americans in response efforts.

There are multiple potential explanations for the above findings. Systemic racism against people of color in the U.S. has led to disparities in housing, wealth, healthcare access, health care quality, and employment.^13,14^ Non-white individuals are more likely to live in more confined housing conditions, be employed as essential workers, and lack health insurance, however race remained an independent predictor of infection after controlling for these factors.^13–15^ Other possible factors that could explain this finding include less access to early testing and a higher likelihood of high viral load due to medical conditions associated with systemic racism and chronic stress, such as asthma, cardiovascular disease, and immunosupression.^16–19^ Further study is necessary to validate these and any other unidentified contributors.

Our INLA models demonstrated that higher population density was positively associated with COVID-19 case burden. This is consistent with existing evidence that COVID-19 transmission may be driven primarily by sustained close contact in confined, unventilated settings.^20^ This is consistent with prior research, however there is also evidence that connectivity between locations may be most critical to spread.^21,22^

It has been widely established that U.S. adults without health insurance have reduced access to high quality care and experience poorer health outcomes.^23^ Our INLA models demonstrated that a higher percentage of uninsured residents in a zip code locality was positively associated with COVID-19 diagnosis. This may be due to delays in isolation or treatment measures due to a reduced quantity of available early testing sites or kits, which has been previously documented in areas of high transmission.^6^ Similarly, patients may decline testing due to fear of associate financial costs. Uninsured individuals may likewise be less likely to afford protective equipment within their households and in public.

Essential service occupations have been linked with higher risk of contracting COVID-19.^24^ Approximately 10% of United States workers are employed in occupations where COVID-19 exposure occurs at least once per week, and 18.4% are employed in occupations where exposure occurs at least once per month.^25^ People of color are more likely to be employed in essential occupations where PPE may be scarce.^26^ Elevated COVID-19 infection risk was found among essential service occupations in a study of Washington State.^27^ We found that service occupation type was not independently associated with risk of infection. It is possible the measure of essential employment excluded pertinent occupations, or was skewed toward a lower-risk essential workplace that was able to ensure social distancing practices and provide PPE. Alternatively, it is possible the effect size of this risk was undetected relative to factors such as race or ethnicity. Further study is necessary to interpret our findings.

Age has been widely studied in its relation to COVID-19 morbidity and mortality, however its role in risk of infection is less well understood.^28^ Outbreaks in some states have been linked to young adults.^29,30^ After accounting for other predictors of infection risk, our model did not find that individuals ages 20-39 contributed significantly to case burden. This may be due to an interaction of factors. Younger people, who present more often with mild disease, may be less likely to seek out or qualify for testing. Alternatively, the distribution of positive cases may be bimodal, with rapid spread throughout nursing homes and long-term care facilities increasing exposure of older adults. The extent to which transmission is propagated by young adults requires further study.

### Methodological Considerations and Limitations

Our sample included ZIP code level data in rural and urban settings and across 19 states. Nonetheless, our sample is limited by the states and municipalities for which data was available at the time of analysis and resource-limited settings may thus be underrepresented. Additionally, differences in the local criteria used by public health agencies to confirm positive cases may have reduced our power to detect associations. Our final analysis included data from 4,803 ZIP codes, but future studies with a wider range of ZIP codes may improve reliability of the associations we report.

### Conclusions

A dynamic evidence base indicates disproportionate risk of COVID-19 infection among some socio-demographic populations.^2,7,14,15,24,25,27^ Our study helps to validate and expand understanding of these inequities, integrating several risk factors into an index reflecting composite vulnerability to infection. Our findings reinforce the urgent need for increased testing accessibility for vulnerable communities at higher risk of infection. Importantly, these results also reinforce the concurrent need to address structural racism and its numerous adverse impacts on the health and healthcare of Black Americans and other communities of color.

## Data Availability

Data is available for public use:

https://chaselab.net/USACovidIndex/ZipIndex.csv

## Notes

### Competing Interest Statement

The authors have declared no competing interest.

### Clinical Trial

Not a clinical trial

### Funding Statement

no external funding was received

### Author Declarations

The study use de-identified publicly available data and did not warrant a review by an Institutional Review Board.

